# Genomic diversity and antimicrobial resistance of *Prevotella* spp. isolated from chronic lung disease airways

**DOI:** 10.1101/2021.08.30.21262864

**Authors:** Kasey A. Webb, Olusola Olagoke, Timothy Baird, Jane Neill, Amy Pham, Timothy J. Wells, Kay A. Ramsay, Scott C. Bell, Derek S. Sarovich, Erin P. Price

## Abstract

Cystic fibrosis (CF) and chronic obstructive pulmonary disease (COPD) are characterised by increasingly frequent acute pulmonary exacerbations that reduce life quality and length. Human airways are home to a rich polymicrobial environment, including members of the obligately anaerobic genus, *Prevotella*. Despite their commonness, surprisingly little is known about the prevalence, role, genomic diversity, and antimicrobial resistance (AMR) potential of *Prevotella* species/strains in healthy and diseased airways. Here, we used comparative genomics to develop a real-time PCR assay to permit rapid *Prevotella* spp. quantification from cultures and clinical specimens. Assay specificity was validated across a panel of *Prevotella* and non-*Prevotella* species, followed by PCR screening of CF and COPD respiratory-derived cultures. Next, 35 PCR-positive isolates were subjected to whole-genome sequencing. Of eight identified species, *P. histicola, P. melaninogenica, P. nanceiensis, P. salivae* and *P. denticola* overlapped between participant cohorts. Phylogenomic analysis revealed considerable interhost but limited intrahost diversity, suggesting patient-specific lineages in the lower airways, probably from oral cavity aspirations. Correlation of phenotypic AMR profiles with AMR gene presence identified excellent correlation between *tetQ* presence and decreased doxycycline susceptibility, and *ermF* presence and decreased azithromycin susceptibility and clindamycin resistance. AMR rates were higher in the CF isolates, reflecting greater antibiotic use in this cohort. All tested *Prevotella* isolates were tobramycin-resistant, providing a potential selection method to improve *Prevotella* culture retrieval rates. Our addition of 35 airway-derived *Prevotella* genomes to public databases will enhance ongoing efforts to unravel the role of this diverse and enigmatic genus in both chronic respiratory diseases and healthy lungs.

**Data summary:** Thirty-five *Prevotella spp*. genomes generated in this study are available in the Sequence Read Archive (SRA) and GenBank databases under BioProject accession PRJNA742126.

## Introduction

Chronic lung diseases are a leading cause of global healthcare burden and early mortality^1,2^. Although cystic fibrosis (CF) and chronic obstructive pulmonary disease (COPD) have fundamentally different aetiologies, both are characterised by impaired airway mucociliary clearance, which provides a favourable environment for microbial colonisation and persistence, and a challenging environment for pathogen eradication. Infections are the primary contributor in CF and COPD respiratory exacerbations (i.e. acute worsening of symptoms), during which inflammation and irreversible airway damage can occur^3^. In CF, 80-95% of deaths are associated with acute exacerbations and chronic bacterial infections, which eventually lead to respiratory failure^4^. In COPD, bacterial, viral, and fungal respiratory infections trigger ∼70-80% exacerbation events^5^. Despite their crucial role in reducing quality and length of life, there remain major gaps in understanding the collective microbial drivers associated with CF and COPD pathophysiology.

Routine culture-based diagnosis of CF and COPD exacerbations often overlooks more fastidious or uncultivable members within the airway microbiome, leaving a gap in fully understanding the complex composition and activity of CF and COPD microbiomes^6,7^. Culture-independent techniques like metataxonomics (e.g. 16S rRNA amplicon sequencing), metagenomics, and metatranscriptomics are gaining traction as adjuncts or potentially even alternatives to culture due to their ability to identify a much broader spectrum of microbes, including uncultured taxa, resulting in better diagnosis, more informed treatment strategies, and improved clinical outcomes^8-10^. In addition to identifying well-characterised microbes associated with respiratory exacerbations, molecular methods have unveiled previously unidentified microbial communities in chronic lung disease airways, including a diverse anaerobic microbiome^11^.

The *Prevotella* genus, which comprises ≥51 species^12,13^, is a common and often abundant obligate anaerobe in the human body, including in CF and COPD airways^11,14-19^. Despite its commonness, the role of *Prevotella* in disease progression is incompletely understood in CF^11^, and even less so in COPD. Several studies investigating *Prevotella* spp. in airway disease, particularly CF, have yielded conflicting evidence. Some studies suggest a likely pathogenic role^18,20^, whereas others have shown no role^21^, or even a benefit when *Prevotella* spp. are abundant^22,23^. A major contributor to these equivocal findings is that culture-dependent and metaxonomic methods largely fail to identify microbes to the species and strain level^7,13,24,25^. As such, only a handful of *Prevotella* studies have described potential species-or strain-specific mechanisms in airway disease pathogenesis.

The number of *Prevotella* genomes has been steadily increasing in recent years. However, in comparison to more intensely studied CF and COPD airway organisms, there is a relative paucity of respiratory *Prevotella* genomic data in public databases. Most *Prevotella* species contain only a single representative (and many were generated from a single metagenomic study)^26^, leading to issues with correct species assignments from metagenome-assembled genomes. Further, most genomic studies have concentrated on known pathogens in oral (e.g. *P. intermedia* and *P. nigrescens*), gut (e.g. *P. copri*), and urogenital (e.g. *P. bivia*) microbiomes^27-29^. There is a lack of knowledge about the *Prevotella* spp. commonly found in airways, particularly in COPD. Insufficient resolution to the species and strain level has limited our understanding of *Prevotella* spp. diversity, function, evolution, and antimicrobial resistance (AMR) potential within CF and COPD airways during exacerbations and stability^13,24^.

Here, we first used a comparative genomics approach to design and optimise a novel real-time PCR assay to permit the rapid, cost-effective, and specific detection of *Prevotella* spp. from both culture and clinical specimens. Next, 43 CF-COPD sputa and bronchial washings were cultured to isolate individual *Prevotella* strains for whole-genome sequencing (WGS) characterisation, followed by genomic analyses to determine species identity, strain relatedness, and within-host evolution. Cultured isolates were subject to AMR testing across 11 clinically-relevant antibiotics, and correlated with AMR determinants identified by WGS. Finally, *Prevotella* morphological characteristics were assessed for their diagnostic value.

## Methods

### Ethics statement

Collection and analysis of clinical samples was approved by The Prince Charles Hospital (TPCH) Human Research Ethics Committee (HREC), project IDs HREC/13/QPCH/127 (CF samples) and HREC/2019/QPCH/48013 (COPD samples). Site-specific approvals were obtained for CF recruitment at TPCH, Brisbane, Australia, and COPD recruitment at Sunshine Coast Hospital and Health Service, Sunshine Coast, Australia. All participants provided written consent.

### Sample collection and storage

A total of 36 CF sputa were obtained from 21 expectorating adults with CF presenting at TPCH Adult CF Centre between 2017 and 2019. Seven participants provided multiple sequential sputa (**Table S1**). Sputa were stored non-anaerobically at -80°C in Luria-Bertani broth containing 20% glycerol, and stored for 2-24 months prior to microbiological assessment. A total of 32 COPD sputa (*n*=21) or bronchial washings (BWs; *n*=11) were obtained from 21 current/former tobacco smokers during community or in-hospital consult at Sunshine Coast Hospital and Health Service in 2020 (**Table S1**). Five participants provided multiple sequential sputa (**Table S1**). BWs were taken from participants with suspected lung cancer, with most collected as paired specimens from both the tumour site (biopsied after BW collection) and approximately the same position in the non-tumour contralateral (contra) lung. BWs were pelleted at 4°C and ∼9,300 x g for 15 min, with supernatant removed and pellet resuspended in 800 µL phosphate-buffered saline. COPD samples were stored within 12-36 hours of collection using the same parameters as described for the CF specimens, and microbiologically processed within ∼2 weeks of collection. Clinical and antibiotic treatment regimens were recorded (**Table S1**) to permit correlation with *Prevotella* AMR profiles.

### Microbiological preparation and storage

Clinical samples were directly subcultured onto anaerobic blood medium containing nalidixic acid and vancomycin (Thermo Fisher Scientific, Seventeen Mile Rocks, QLD, Australia; cat. no. PP2065). Plates were incubated anaerobically at 37°C in an AnaeroPack 2.5L jar with an AnaeroGen 2.5L gas pack (Thermo Fisher) for 2-5 days. Isolates were subcultured based on different morphotypes, with a second subculture for purity onto non-selective anaerobic blood medium (Thermo Fisher; cat. no. PP2039). *Prevotella* real-time PCR-positive isolates were again sub-cultured onto non-selective anaerobic blood medium for purity and stored non-anaerobically at -80°C in 1mL Luria-Bertani broth containing 20% glycerol. Additionally, WGS-confirmed, representative *Prevotella* isolates derived from CF samples were cultured from glycerol stocks onto both polyvitaminic-supplemented chocolate agar (PVCA; Edwards Group, Murrarie, QLD, Australia; cat. no. 1065) and non-selective anaerobic blood medium to assess phenotypic differences over a 10-day period.

### DNA extractions

DNA of presumptive *Prevotella* spp. isolates was initially extracted using 5% chelex-100^30^ (Bio-Rad, Gladesville, NSW, Australia), heat-treated at 95°C for 20 min, and diluted to 1:50 with molecular-grade H_2_O prior to PCR. DNA for WGS was extracted using the Gram-positive bacteria protocol of the DNeasy Blood and Tissue Kit (Qiagen, Chadstone, VIC, Australia). Qiagen extractions were quality-checked and quantified using the NanoDrop 2000 spectrophotometer (Thermo Fisher).

Clinical specimens were extracted using 3 parts TRI Reagent LS (Sigma-Aldrich, North Ryde, NSW, Australia) to 1 part specimen, 30µL 0.1M 2-mercaptoethanol, and ∼ 100µL equal mixture of 0.1 and 0.5mm zirconia beads (Daintree Scientific, St Helens, TAS, Australia). Homogenisation comprised eight rounds of 30 sec bead beating (each round followed by 30-60 sec on wet ice) using the ‘medium’ setting of the Minilys tissue homogeniser (Bertin Instruments, Montigny-le-Bretonneux, France). After removal of the RNA aqueous phase, back extraction was performed on the interphase and proteinaceous phase^31^ to isolate total DNA.

### *Prevotella* spp. real-time PCR assay design

Due to high levels of genetic variability among *Prevotella* spp., the highly conserved 16S-23S rRNA genes were targeted. A custom BLAST^32^ 2.10.2+ database was created using 60 *Prevotella* spp. genomes encompassing 26 assigned and nine unassigned species (**Table S2**). Discontiguous MegaBLAST was carried out against the 4.8kb *P. intermedia* ATCC 25611 23S-16S rRNA locus (GenBank accession CP019300.1, coordinates 63001-67800) using the custom BLAST database. BLAST .xml outputs were aligned and visualised using Geneious Prime vR10. A highly conserved area within the *Prevotella* spp. 16S rRNA gene was assessed further to identify potential PCR oligos, with conserved regions possessing several single nucleotide polymorphisms (SNPs) in other microbes exploited during probe and primer design. Primers and probes were initially assessed using Primer Express v3.0 (Applied Biosystems) to test melting temperature (*T*_m_) suitability in Black Hole Quencher format (primer *T*_m_=60°C; probe *T*_m_=70°C). All candidate primers and probes were assessed for self- and hetero-dimer formation using Beacon Designer (http://www.premierbiosoft.com/qpcr/), with oligo sequences requiring a ΔG value of >-4 kcal/mol. Candidate oligos were subsequently subjected to NCBI Microbial Nucleotide BLAST and Nucleotide BLAST searches to confirm specificity for *Prevotella* spp.. As the 16S region is highly conserved in all bacteria, the forward primer incorporated a penultimate mismatch to prevent non-specific binding to closely related non-*Prevotella* taxa. Additionally, a BHQplus probe (CAL Fluor Gold 540-pdCApdUApdUpdCGApdCGGGGpgUGG-BHQ1; Biosearch Technologies, Petaluma, California, USA) was included to enhance specificity and to permit assay use on clinical specimens. Forward (5’-TTCTCCAGtCCAGGATGTGtC-3’) and reverse (5’-CGGGGATAACAGGCTGATCC-3’) primers were manufactured by Macrogen Inc. (Geumcheon-gu, Seoul, South Korea), with lowercase nucleotides representing intentional mismatches.

Real-time PCRs were carried out on a Bio-Rad CFX96 thermocycler using 0.35µM primers and probe, 1X Sso Advanced Universal mastermix (Bio-Rad), molecular-grade H2O, and 1µL template, to a 5µL final volume. Thermocycling parameters were: 95°C for 2 min, followed by 45 cycles of 95°C for 5 sec and 60°C for 20 sec. DNA integrity for all clinical samples was assessed via amplification towards β-globin^33^ (human) and universal 16S rRNA^34^ (bacteria); the universal 16S rRNA assay was also used to verify bacterial culture DNA integrity, and a panfungal assay (O. Olagoke and E. Price, unpublished) was used to verify *Candida* spp. DNA integrity.

### Real-time PCR assay optimisation and specificity testing

The final optimised real-time PCR assay was assessed for accuracy by running against a diversity panel of six *Prevotella* spp. (one each of *P. copri, P. denticola, P. histicola, P. melaninogenica, P. nigrescens*, and *P. salivae*), 1 human DNA sample, and 40 non-*Prevotella* spp., with a focus on common respiratory tract microbes (*n*=1 unless otherwise stated): *Achromobacter xylosoxidans, Bordetella bronchiseptica, Burkholderia cepacia, Burkholderia territorii, Burkholderia thailandensis, Candida albicans, Candida dubliniensis, Cupriavidus metallidurans, Enterobacter aerogenes, Enterobacter cloacae* (*n*=3), *Enterococcus raffinosus, Klebsiella oxytoca, Klebsiella pneumoniae* (*n*=2), *Lacticaseibacillus paracasei* (*n*=2), *Pseudomonas aeruginosa* (*n*=12), *Staphylococcus aureus* (*n*=3), *Staphylococcus haemolyticus, Stenotrophomonas maltophilia, Veillonella atypica* (*n=*3), *Veillonella parvula*, and *Veilonella dispar*. All DNA was normalised spectrophotometrically to 2ng/µL prior to PCR.

Limits of detection (LoD) and quantification (LoQ) for the optimised assay were determined using *P. melaninogenica* LMG 28911 DNA, using serial dilutions of 4ng/µL to 4 × 10^−6^ ng/µL across 8 replicates per dilution. The upper and lower LoD/LoQ limits were determined as described elsewhere^35^. Genome equivalence was determined based on an average *Prevotella* genome size of 3.29Mbp and a single copy of the 16S rRNA gene.

### 16S rRNA gene diversity profiling (metataxonomics)

Metataxonomics was performed by the Australian Genome Research Facility (AGRF; Melbourne, VIC, Australia) for total CF DNA and ID Genomics (Seattle, WA, USA) for total COPD DNA using standard protocols for V3-V4 (i.e. 341F and 806R) primers.

### Antimicrobial resistance (AMR) profiling

Twenty-eight glycerol-stocked *Prevotella* cultures were subcultured onto non-selective anaerobic blood medium, or PVCA where no growth was obtained on the anaerobic medium (*n*=2), and incubated at 37°C under anaerobic conditions for 2-3 days. Cultures were then resuspended into phosphate-buffered saline to OD_600_=0.75 (Biochrom, Cambridge, UK). Six supplemented chocolate plates per isolate were lawn-inoculated with the OD-normalised cells as previously described^36^, and antibiotic discs (Edwards Group) towards 11 antibiotics (amoxicillin-clavulanate [AMC] 30µg; azithromycin [AZM] 15ug; ceftazidime [CAZ] 30µg; chloramphenicol [CHL] 30µg; clindamycin [CLI] 2µg; doxycycline [DOX] 30µg; imipenem [IPM] 30µg; meropenem [MEM] 10µg; metronidazole [MZ] 5µg; piperacillin-tazobactam [TZP] P:100µg, TZ:10µg; tobramycin [TOB] 10µg) were placed onto inoculated plates. Plates were incubated at 37°C under anaerobic conditions for 48h prior to measurement. As no disc diffusion reference ranges exist for *Prevotella* spp., growth up to the disc (i.e. 6mm diameter) was denoted as AMR, which underestimates the true rate of AMR but provides a clear cut-off for unambiguous interpretation. Diameters ≤30mm for any antibiotic were denoted as reduced susceptibility.

### Whole-Genome Sequencing (WGS)

PCR-confirmed *Prevotella* isolates were sequenced using the Illumina HiSeq 2500 platform with 150bp seqWell plexWell paired-end read chemistry at AGRF.

### Genome assemblies

*Prevotella* genomes were *de novo* assembled using default Microbial Genome Assembly Pipeline v1.1 parameters (https://github.com/dsarov/MGAP---Microbial-Genome-Assembler-Pipeline) or SPAdes^37^.

### Speciation and mixture analysis

*Prevotella* spp. assemblies were initially assessed for species identification and mixtures using the Ribosomal MLST^38^ tool. Isolates containing mixtures of *Prevotella* and non-*Prevotella* species (*n*=8), or multiple *Prevotella* species (*n*=1) (**Table S3**), were excluded from further investigation.

### Phylogenetic analysis

A custom BLAST database of *Prevotella* genomes (*n*=234) comprising taxa of known (*n*=56) and unassigned (*n*=40) species, and including non-mixed isolate genomes sequenced in this study (*n*=35 genomes), was created using BLAST 2.10.2+. BLAST analysis was performed against the full-length 1,534bp *P. melaninogenica* 16S rRNA gene (RefSeq ID: NC_014371.1; bases 962,784-964,317). Hits were filtered to remove redundancies (final *n*=199), and reads aligned using ClustalX2 v2.1^39^. A dendogram was constructed using the neighbour-joining method and the Tamura-Nei model and viewed in FigTree v1.4.0 (http://tree.bio.ed.ac.uk/software/figtree/) using midpoint rooting.

### Phylogenomics

Variant identification of our CF-COPD genomes (‘SCHI’ prefix) was performed using SPANDx v4.0^40^. Reference genomes for *P. denticola, P. histicola, P. melaninogenica, P. nanceiensis*, and *P. salivae* were downloaded from GenBank (IDs GCA_000193395.1, GCA_000234055.1, GCA_000144405.1, GCA_000163055.2, and GCA_000477535.1, respectively) and used for read mapping. Synthetic reads were generated from GenBank genome assemblies for *P. denticola* (GCA_000191765.2, GCA_000759205.1, GCA_000421205.1, GCA_003515045.1, and GCA_900454835.1), *P. histicola* (GCA_000613925.1), *P. melaninogenica* (GCA_000163035.1, GCA_000759305.1, GCA_002208725.2, GCA_003609775.1, and GCF_000517825.1), *P. nanceiensis* (GCA_000517825.1 and GCA_000613985.1), and *P. salivae* (GCA_000185845.1 and GCA_902374235.1) using ART v2.3.7^41^.

Illumina reads for *P. histicola* T05-04T (SRR1518629) and *P. nanceiensis* ADL-403 (SRR755359) were downloaded from the Sequence Read Archive database and quality filtered using Trimmomatic v0.39^42^ parameters described elsewhere^43^. The resulting biallelic, orthologous SNP matrix generated by SPANDx was phylogenetically reconstructed using the maximum parsimony heuristic function of PAUP* v4.0a (build 168). Phylogenies were visualised in FigTree; all trees were midpoint-rooted.

### AMR determinant analysis

The Resistance Gene Identifier v5.1.1, which employs the Comprehensive Antibiotic Resistance Database (CARD) v3.1.0^44^, was used to identify putative AMR determinants.

## Results and Discussion

Molecular typing approaches have been instrumental in unveiling previously untapped microbial diversity in both healthy and diseased lungs. Using these techniques, *Prevotella* spp. have consistently been identified as one of the most prevalent taxa in CF and COPD airways^10,14,19^. However, most studies to date have employed 16S metataxonomics (usually targeting the V3-V4 gene region), which only characterises *Prevotella* to the genus level, despite species and strain resolution being fundamental to unravelling the role of these anaerobes in chronic lung disease. To address this knowledge gap, our study used a combination of culture-dependent and -independent methods to examine the genomic and phenotypic diversity, and prevalence, of *Prevotella* spp. present in CF and COPD airways.

We first used a comparative genomics-guided approach to develop an accurate real-time PCR assay to enable the rapid detection of *Prevotella* spp., to the exclusion of all other organisms, both from cultures and clinical specimens. Our PCR assay was reasonably sensitive, with LoD and LoQ both being 400fg/µL, or 111 genome equivalents (**Figure 1**). When tested against a diverse panel of 47 microbial and human DNA, the PCR demonstrated 100% specificity, amplifying all six tested *Prevotella* species, but none of the non-*Prevotella* spp. DNA and 10 no-template controls. On clinical specimens, the PCR identified *Prevotella* in 81% CF and 71% COPD participants, and in 81% CF and 66% COPD specimens (**Table 1**). This difference in detection rates between diseases was expected given the generally lower total bacterial load in COPD airways^1^. Metataxonomics had higher *Prevotella* sensitivity than PCR, identifying *Prevotella* in six PCR-negative samples, although there was one false-negative (SCHI0049.C.3, which was PCR-and culture-positive); there was otherwise near-perfect correlation between PCR and metataxonomics (**Table S3**). In the six PCR-negative samples, *Prevotella* relative abundance was very low (0.004-0.3%; median=0.05%), indicating that DNA levels were below the PCR LoD threshold. Thus, culture-independent methods are highly suitable for accurate *Prevotella* spp. detection from polymicrobial specimens, particularly real-time PCR, which is often performed upon clinical sample receipt, thereby permitting same-day detection of *Prevotella* in respiratory samples.

**Figure 1.**
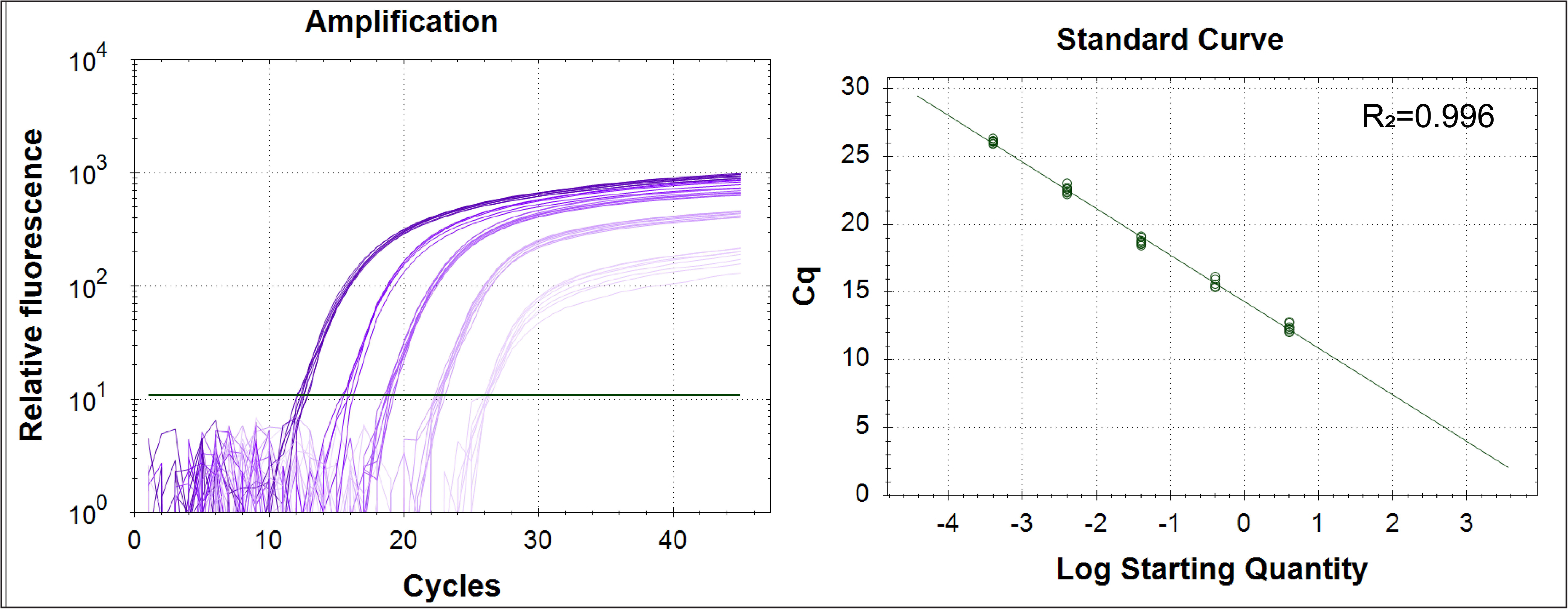
Limits of detection (LoD) and quantification (LoQ) for the *Prevotella* real-time PCR assay. The LoQ and LoD were both 400fg/µL according to previously defined parameters^35^, or 111 genome equivalents.

**Figure 2.**
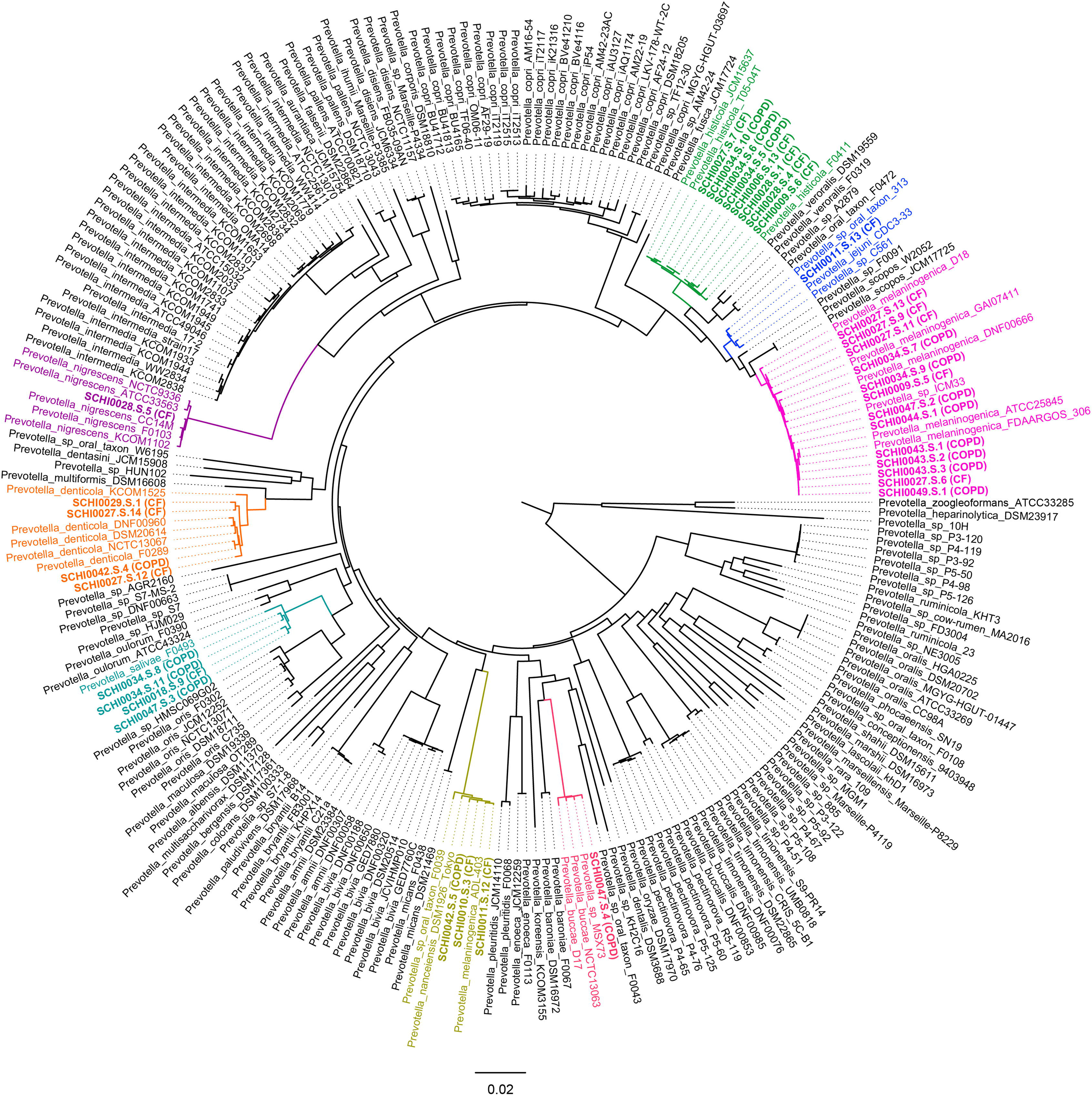
Full-length 16S ribosomal RNA gene phylogenetic analysis of the 35 *Prevotella* strains retrieved in this study (identified by ‘SCHI’ prefix) against 199 non-redundant public *Prevotella* genomes comprising 56 taxa of known species and 40 taxa of unassigned species. Species clades containing isolates identified from our cystic fibrosis (CF) and chronic obstructive pulmonary disease (COPD) specimens are coloured.

**Table 1.** *Prevotella* spp. prevalence in cystic fibrosis (CF) and chronic obstructive pulmonary disease (COPD) according to microbiological culture, real-time PCR, and 16S rRNA gene metataxonomics.

| *Prevotella* +ve |  | CF | COPD |
| --- | --- | --- | --- |
| Participants | Culture | 11/21 (52%) | 6/21 (29%) |
|  | PCR | 17/21 (81%) | 15/21 (71%) |
|  | Metataxonomics | 15/17 (88%) | 16/18 (89%) |
| Specimens | Culture | 16/36 (44%) | 9/32 (28%) |
|  | PCR | 29/36 (81%) | 21/32 (66%) |
|  | Metataxonomics | 26/31 (84%) | 20/22 (91%) |

Culture of sputa and BWs onto three different agars (anaerobic blood medium containing nalidixic acid and vancomycin, non-selective anaerobic blood medium, and PVCA) yielded a much lower *Prevotella* positivity rate compared with PCR and metataxonomics. Only 44% CF and 28% COPD clinical specimens, respectively, were *Prevotella*-positive via culture, compared with 81% and 66% using PCR, and 84% and 91% using metataxonomics (**Table 1**). The reduced retrieval rate in COPD vs. CF can be explained by lower relative *Prevotella* abundance according to metataxonomics (0.3-74.3% [median=9.6%) for CF vs. 0.002%-36.4% [median=0.6%] for COPD). The lower prevalence of *Prevotella* using culture is likely due to multiple factors, such as an inability of culture-independent methods to distinguish viable from non-viable cells, nutritional fastidiousness, cells being in a viable but non-culturable state, and outcompetition from faster-growing taxa^9,19,22^. A prior study found that four different culture media were required to recover 42 *Prevotella* isolates from 26 samples, and only 20 isolates were recoverable on more than one medium^19^. Moreover, all four media were required to recover *P. melaninogenica* from all sputum samples^19^, suggesting that growth requirements may be species- and strain-specific. In contrast to this prior work, we used aerobic collection and storage conditions, which likely precluded recovery of *Prevotella* taxa with poor/no aerotolerance^45^. Consistent with their fastidiousness, one *P. histicola* and five *P. melaninogenica* initially isolated in our study were unable to be resuscitated from glycerol stocks upon repeated attempts, despite previously being culturable. Taken together, it is probable that several *Prevotella* species or strains were not recovered by culture in our study due to multiple factors, thereby underestimating species and strain diversity within our cohorts.

To gain greater insight into *Prevotella* residing in CF and COPD airways, we performed WGS and phylogenetic analysis on cultured isolates to determine species identity and strain relatedness. From 36 CF sputa, 27 PCR-positive *Prevotella* colonies were retrieved, of which 18 were *Prevotella* spp.; the remaining nine contained a mixture of *Prevotella* and other taxa, or a mixture of multiple *Prevotella* spp., due to insufficient subculturing for purity prior to WGS (**Table S3**). To avoid known bioinformatic analysis issues associated with variant calling from mixtures^46^, these mixed data were excluded from phylogenetic analysis. Among the 18 isolates, WGS identified seven *Prevotella* spp.: *P. histicola* (*n*=5), *P. melaninogenica* (*n*=5), *P. denticola* (*n*=3), *P. nanceiensis* (*n*=2), *P. jejuni* (*n*=1), *P. nigrescens* (*n*=1), and *P. salivae* (*n*=1). Of the 17 COPD-derived isolates, none were mixed due to a greater number of passages carried out to obtain purity. WGS identified six *Prevotella* spp.: *P. melaninogenica* (*n*=8), *P. histicola* (*n*=3), *P. salivae* (*n*=3), *P. buccae* (*n*=1), *P. denticola* (*n*=1), and *P. nanceiensis* (*n*=1) (**Table S3**). Despite being fundamentally different diseases, there was little difference in *Prevotella* species found in CF and COPD, with only *P. jejuni* and *P. nigrescens* being unique to CF, and *P. buccae* being unique to COPD. One explanation for this overlap is that the oral cavity and oropharynx are common habitats for all retrieved species, except *P. jejuni*^47^; it is thus possible that some strains we isolated were upper airway residents. Whilst oral cavity contamination is unavoidable during respiratory secretion collection, especially for sputum, microbes can also enter lower airways via small-volume oral cavity aspirations; indeed, the risk of microbial colonisation and persistence is greatly increased in mucociliary-impaired CF and COPD airways^24,48-52^. In support of lower airway origin, we identified *Prevotella* spp. in both sputa and BWs. Unlike sputa, BWs are collected via bronchoscopy and thus far less prone to oral/oropharynx contamination, providing greater confidence that many of our retrieved *Prevotella* isolates originated from the lower airways.

The origin of *P. jejuni* in one CF participant is somewhat perplexing. This species was first documented in the small intestine of a child with coeliac disease in 2012^53^, and little is currently known about the distribution of this organism in healthy and diseased niches. One possibility is that *P. jejuni* entered the CF airways via inhalation of gastroesophageal aspirations. Alternatively, microbial inter-compartment cross-talk between the lungs and gut (the ‘Gut-Lung axis’) via the mesenteric lymphatic system may explain its presence in the lungs^54^. As a relatively new taxon, little is known about possible mechanisms of *P. jejuni* pathogenesis, with more work needed to determine the prevalence and virulence potential of this bacterium in both healthy and diseased airways.

The most frequently isolated species in our study – *P. melaninogenica* (37%), *P. histicola* (23%), *P. salivae* (11%), *P. denticola* (11%), and *P. nanceiensis* (9%) – are common in CF^14,15,19^ and COPD^25^ airways. However, their precise role in airway disease is poorly understood due to conflicting reports about their beneficial vs. pathogenic potential. For instance, *P. melaninogenica, P. salivae*, and *P. nanceiensis* may inhibit non-typeable *Haemophilus influenzae*-induced pro-inflammatory cytokine production by lung dendritic cells^52^, increased *P. melaninogenica* prevalence in CF airways has been associated with improved lung function^15^, and *P. histicola* can mute pro-inflammatory pathways triggered by *P. aeruginosa* in CF airways^55^. In contrast, *P. melaninogenica* produced the greatest variety of short-chain fatty acids (associated with inflammation) amongst five tested anaerobes^18^, and *P. denticola* shares similarities in its lipid A moieties to *P. intermedia* (a known pathogenic species), pointing towards pro-inflammatory traits^56^. Finally, certain strains of the periodontal disease-associated species, *P. nigrescens*, a close relative of *P. intermedia*, are thought to encode an enzymatic cell hydrolysis mechanism that destroys tissues. Given the reported conflicting roles of *Prevotella* spp. in chronic lung disease, these collective results suggest that virulence mechanisms vary among *Prevotella* strains, which requires further exploration^57,58^. Our genomic data will assist these future efforts.

In our experience, metagenomic analysis tools can struggle with *Prevotella* species-level identification from respiratory specimens due to a lack of accurate representative species in public databases. Exemplifying this problem, few publicly available genomic data are available for *P. nanceiensis* – only three were available at the time this study was undertaken, one of which is misclassified as *P. melaninogenica*, and another as *Prevotella* sp. (**Figure 3**). Thus, our addition of three *P. nanceiensis* genomes doubles the available *P. nanceiensis* data. Similarly, there are currently only a handful of nonredundant *P. jejuni* genomes in public databases, and none are from lower airway specimens; our addition *P. jejuni* SCHI0011.S.13 genome is therefore valuable for future respiratory and gastroenterological studies. Taken together, our *P. nanceiensis* and *P. jejuni* genomes will help public database curation efforts, and in turn, improve species identification using metagenomics.

**Figure 3.**
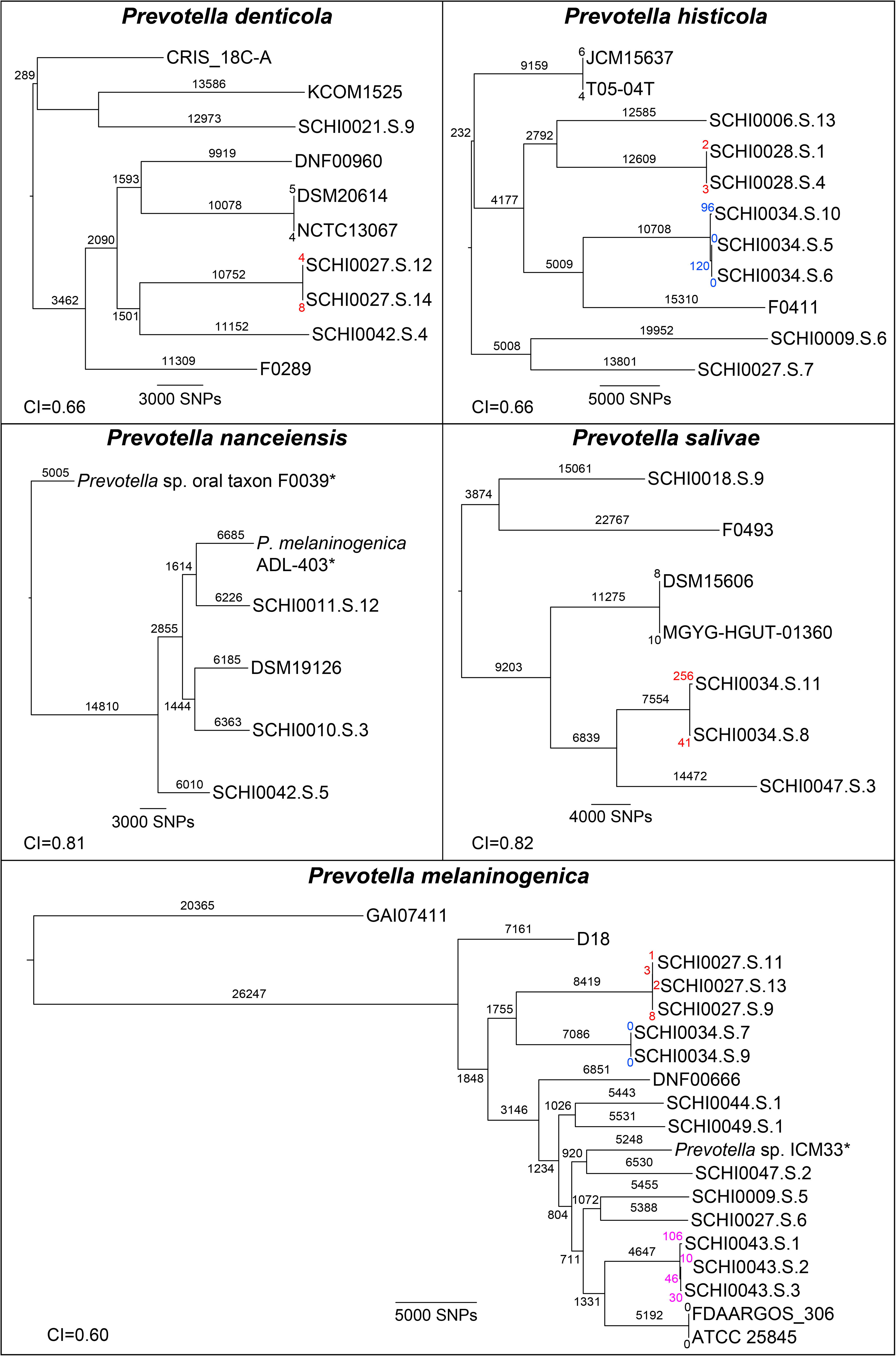
Maximum parsimony within-species phylogenomic reconstruction of *P. denticola, P. histicola, P. melaninogenica, P. nanceiensis* and *P. salivae*. Strains identified from COPD and CF specimens from this study are labelled with the ‘SCHI’ prefix; the remaining strains are public genomes. The number of SNPs separating each strain is labelled on branches; where two or more strains were retrieved from a single participant, coloured branch labels represent the number of SNPs separating these strains. CI, consistency index. Incorrectly speciated public genomes are denoted by an asterisk.

To determine strain relatedness within and between participant airways, we undertook within-species analyses of our *P. denticola, P. histicola, P. melaninogenica, P. nanceiensis* and *P. salivae* genomes (**Figure 3**). Whole-genome SNP analysis revealed high levels of strain and interhost diversity in all examined *Prevotella* species. For example, genetic variation was as high as ∼72k SNPs among *P. histicola* strains, an impressive feat for an organism encoding a 3Mbp genome, and *P. nanceiensis* strains SCHI0010.S.3 and SCHI0011.S.12 were more closely related to strains isolated from France and the US, respectively, than to other strains retrieved from other Australian participants. In contrast, intrahost diversity was relatively limited (0-297 SNPs), with two instances of genetically indistinguishable strains: *P. histicola* SCHI0034.S.5 and SCHI0034.S.6, and *P. melaninogenica* SCHI0034.S.7 and SCHI0034.S.9 (**Figure 3**); the latter pair were retrieved from a BW and sputum collected on the same day (**Table S1; Table S3**). The only exception was *P. melaninogenica* from SCHI0027, where two genetically distinct lineages separated by >20k SNPs were retrieved from this participant. There was no evidence of genetically-related strains between any of the Australian participants for any *Prevotella* spp. (**Figure 3**). These findings indicate that interhost diversity is very high, whereas within-host *Prevotella* diversity is limited, possibly due to frequent microaspiration of *Prevotella* from upper to lower airways (or via the gut-lung axis) rather than chronic infection; however, more in-depth sampling is required to investigate *Prevotella* persistence.

Antibiotics are routinely used in the maintenance and treatment of acute respiratory infections in CF and COPD^15,59^. Given its uncertain pathogenic role, *Prevotella* spp. are not targeted by these treatment regimens^60^. Despite this, AMR rates in *Prevotella* are trending upwards^59^, possibly due to bystander selection. To examine this phenomenon, we determined AMR profiles for all culturable *Prevotella* strains (*n*=27) using disc diffusion against 11 antibiotics commonly used to treat lung infections^15,59^, or which have known activity against anaerobic taxa^47^. AMR (AZM, CAZ, CLI, MZ, TOB) or reduced susceptibility (AMC, AZM, CAZ, DOX, MEM) was observed in at least one strain, whereas all were susceptible towards CHL, TZP, and IPM, an observation supported by previous studies^47,59,61^. Reflecting much greater lifetime antimicrobial use in the CF cohort^62^, the 18 tested CF strains had much higher AMR incidence (57% vs. 8%) and AMR gene presence (83% vs. 35%) compared with COPD strains, and this rate increased to 100% (69% for COPD) when including strains with reduced susceptibility towards one or more antibiotics (**Table 2**). In addition, almost all CF participants were undergoing antimicrobial treatment with at least two different antibiotic classes at the time of sample collection, vs. almost no antibiotic use in the COPD participants (**Table S1**). One strain, CF isolate *P. histicola* SCHI0009.S.6, could be classed as multidrug-resistant due to exhibiting AMR towards three antibiotic classes^63^ – β-lactams, lincosamides, and imidazoles – along with reduced susceptibility towards the tetracycline-class antibiotic, DOX (**Table 2**). Our findings confirm bystander AMR selection in off-target *Prevotella* as a direct result of heavy antimicrobial use in CF (and to a much lesser extent, COPD), which may have ramifications for *Prevotella* persistence as a potential driver of disease pathogenesis.

**Table 2.**
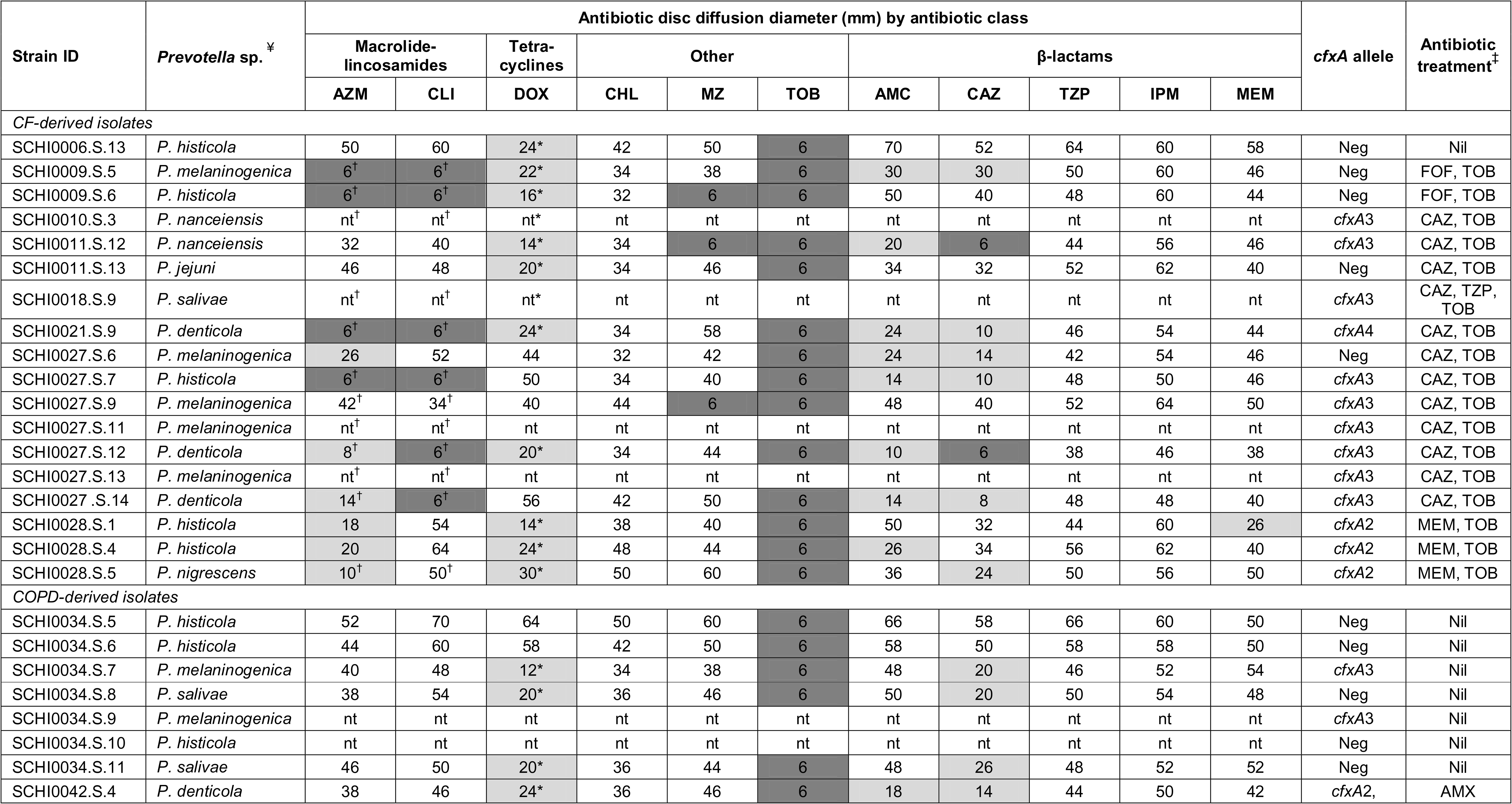
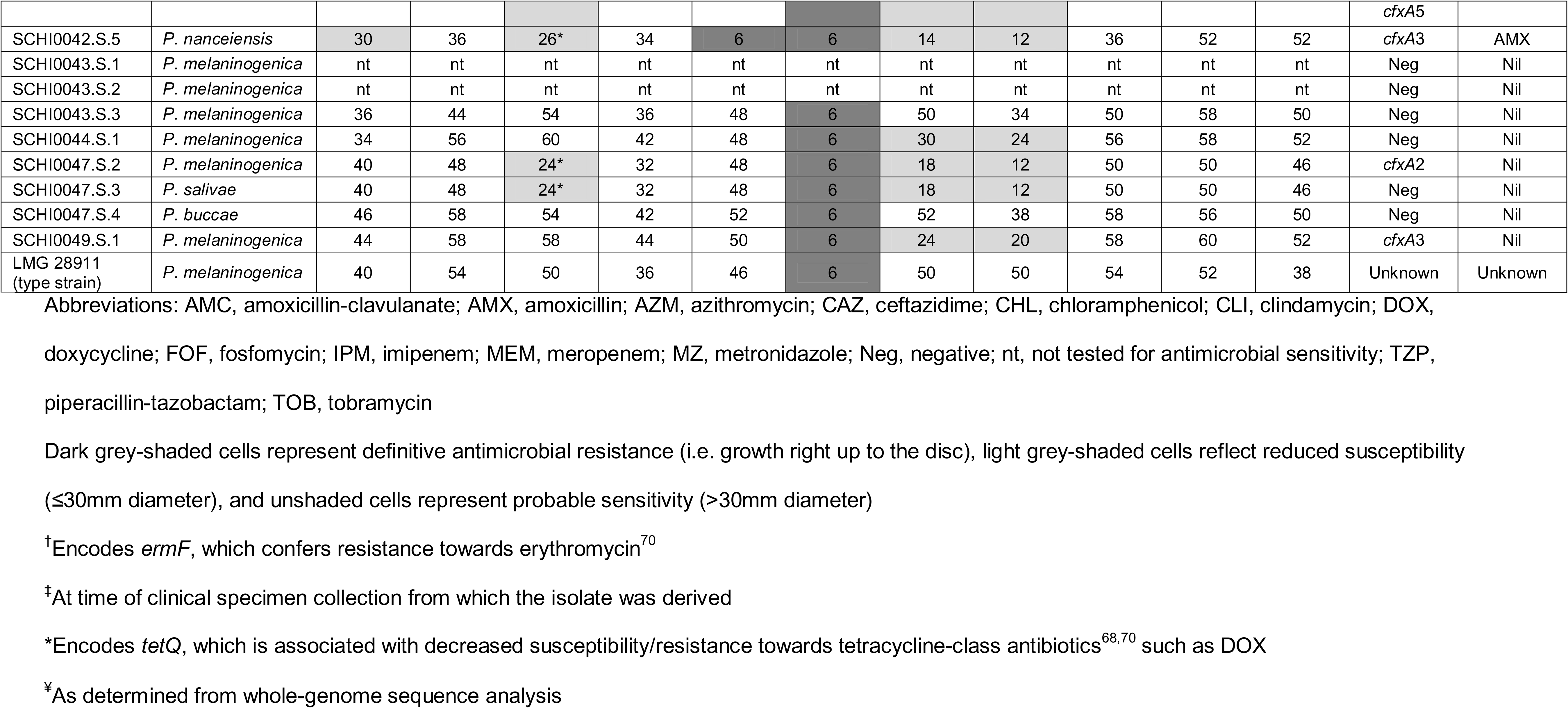
Disc diffusion and antimicrobial resistance gene results for genome-sequenced *Prevotella* isolates from cystic fibrosis (CF) or chronic obstructive pulmonary disease (COPD) respiratory secretions, and antibiotic treatment at time of isolate collection.

TOB was the only antibiotic tested that showed AMR in all tested isolates, including the type strain, *P. melaninogenica* LMG 28911 (**Table 2**). This finding that concurs with prior work reporting 100% TOB resistance among 157 *Prevotella* isolates^59^. A CF microbiome study also found that *Prevotella* abundance was largely unchanged upon TOB treatment^64^. These collective findings indicate that all respiratory *Prevotella* species are intrinsically resistant to TOB. Notably, we observed that the median relative *Prevotella* abundance according to metataxonomics was quite different between CF (9.6%) vs. COPD (0.6%) specimens (**Table S3**). Although the basis of this higher relative *Prevotella* abundance in CF is unknown, one possibility is that most of our CF participants were receiving TOB at the time of sample collection (**Table S1**; **Table 2**), whereas no COPD participant was receiving this antibiotic. The predominance of *Prevotella* spp. in the CF airways might thus be attributed to much more frequent TOB use in this cohort^65-67^. Although not assessed in this study, adding TOB to selective media may improve *Prevotella* recovery rates from respiratory specimens by suppressing growth of other taxa. After TOB, CLI resistance was most common (6/14 CF isolates), followed by AZM (4/14 CF isolates), MZ (1/13 COPD and 3/14 CF isolates), and CAZ (2/14 CF isolates). Decreased susceptibility towards AMC, CAZ, and DOX was common in both CF and COPD isolates; however, only CF isolates exhibited decreased AZM susceptibility (6/14 isolates). These results provide further evidence of bystander selection in *Prevotella* spp., with this effect especially noticeable in CF-derived isolates.

AMR genes represent a major concern in infection control as these genes can rapidly transfer AMR among different genera and species. To determine AMR gene potential, CARD analysis was performed on the 35 CF-COPD *Prevotella* genomes and used to correlate horizontally acquired AMR-conferring genes with AMR/decreased susceptibility phenotypes. Consistent with their higher AMR rates, CF isolates demonstrated greater prevalence of AMR-conferring genes compared with COPD (**Table 2**). Strikingly, 67% CF strains encoded *ermF*, a horizontally-acquired gene that confers resistance towards macrolide, lincosamide, and streptogramin-class antibiotics such as AZM and CLI^47,61,68^. In contrast, *ermF* was absent in all COPD strains, a somewhat unexpected finding given that COPD participants are frequently given long-term macrolides (e.g. azithromycin, erythromycin) as prophylaxis to reduce exacerbation frequency^69^. None of the CF or COPD participants were receiving these antibiotics at the time of sample collection, suggesting that cross-resistance or prior CF antibiotic treatment may be driving *ermF* acquisition and maintenance in *Prevotella. ermF* presence correlated well with AZM and CLI AMR/decreased susceptibility (87% and 75%, respectively). *ermF* rates in our Australian isolates exceed those previously reported (23-68% in UK, USA, and Italian isolates from both healthy and diseased participants)^68,70^. This discrepancy is likely due to only respiratory disease-associated isolates being tested in our study, but may also be due to differences in AMR cut-offs between studies, *in vivo* antibiotic exposure levels, and potentially species or strain composition, noting that one study only identified most isolates to the genus level^68^.

*tetQ*, which confers resistance to tetracycline-class antibiotics (e.g. DOX)^68,70^, was present in 54% of our strains, and more common in CF-derived strains (67% vs. 41%). This rate is much higher than *tetQ* prevalence reported in UK CF (21%)^68^ and human-derived Dutch (30%)^61^ strains.. Our *tetQ*-harbouring strains consistently showed much smaller DOX disc diameters (12-30mm; median 22mm) compared with *tetQ-*negative strains (40-64mm; median 54mm) with 100% correlation between *tetQ* presence and decreased DOX susceptibility (**Table 2**). A prior study reported that 83% of *tetQ*-positive isolates demonstrated intermediate or full resistance towards tetracycline^68^, suggesting that *tetQ* is a reliable marker of reduced susceptibility towards tetracycline-class antibiotics. Taken together, these results confirm that *tetQ* presence robustly confers decreased DOX susceptibility in *Prevotella* spp. Although our dataset is small, we propose *Prevotella* spp. DOX disc diameters of ≥40mm to denote strains lacking *tetQ*, and ≤30mm to denote strains that may be harbouring *tetQ*, with the caveat that other enigmatic determinants conferring reduced DOX susceptibility (e.g. chromosomal mutations) may be present in lieu of *tetQ*. These criteria also differentiate *Prevotella* spp. exhibiting decreased DOX susceptibility from DOX-sensitive strains.

The *cfxA*-type β-lactamase gene is the most clinically important AMR mechanism in *Prevotella* spp.^71^ as it confers AMR towards cephamycins^61^ and some other β-lactam antibiotics^68^, which are commonly used in respiratory infection treatment^59^. Concerningly, *cfxA* is often encoded on mobile elements, meaning that AMR can potentially be transmitted from *Prevotella* spp. to known pathogenic taxa in the respiratory microbiome^72^. We found *cfxA* alleles 2, 3, 4, and/or 5 in 54% strains (72% CF and 35% COPD). *cfxA*3 and *cfxA*2 were most common, being present in 37% and 14% strains, respectively. In comparison, a study of UK and US isolates reported much higher *cfxA* carriage rates (86%), with 29% and 52% strains encoding *cfxA*3 and *cfxA*2, respectively. Their study also found that *cfxA*3-encoding isolates exhibited significantly higher CAZ resistance levels^68^, which we also observed in our *cfxA*3-harbouring strains (median disc diameter=12mm vs. 32mm in non-*cfxA*3 strains) (**Table 2**). In *cfxA*3-encoding strains displaying CAZ sensitivity, reduced *cfxA*3 expression^73^ may explain this phenotype, although this was not explored in our study. In addition, we did not test *Prevotella* susceptibility towards cephamycins so were unable to determine whether *cfxA* variants correlated with these β-lactam antibiotics.

MZ resistance was evident in 4/28 tested strains (3 CF; 1 COPD), despite none of the participants receiving nitroimidazoles at the time of sample collection. Other than TOB, MZ was the only antibiotic that exhibited AMR in any of the COPD strains. The *nim* genes, which encode for nitroimidazole reductase enzymes and have previously been associated with MZ resistance in *P. baroniae*^74^ and *P. bivia*^75^, were not detected in any isolate. Therefore, the basis of MZ resistance in our isolates remains unknown.

Of the eight species identified in our study, four are conventionally considered dark-pigmented – *P. denticola, P. histicola, P. melaninogenica*, and *P. nigrescens*. Pigmented *Prevotella* reportedly harbour a greater cache of virulence factors and are more commonly associated with infection^47^. However, we found limited phenotypic differences within and between species on non-selective anaerobic blood medium at Day 3 (**Figure 4**), with most presenting as convex, shiny, white colonies; only *P. nigrescens* demonstrated tan pigmentation in the primary streak at this time point (**Figure 4F**). In contrast, a dark brown to black pigment was produced by *P. nigrescens* on PVCA by Day 3 (**Figure 5D**), but not on selective or non-selective anaerobic media, even by Day 10, with only medium-brown colonies present (**Figure 5B and 5C**). *P. histicola* demonstrated tan-coloured pigmentation on both non-selective anaerobic medium and PVCA, but only after five days (**Figure 6**), and unlike *P. nigrescens*, did not develop black pigment on PVCA at Days 5 or 10. *P. melaninogenica* reportedly produces a characteristic dark pigment that is commonly associated with the species^76,77^; yet, no pigmentation was observed in the six *P. melaninogenica* isolates on any medium by Day 10. A likely explanation for this phenomenon is that different strains require different media for pigment production^78^. Collectively, the limited pigmentation observed among *Prevotella* spp. suggests that phenotypic characteristics alone should not be used for *Prevotella* genus or species identification.

**Figure 4.**
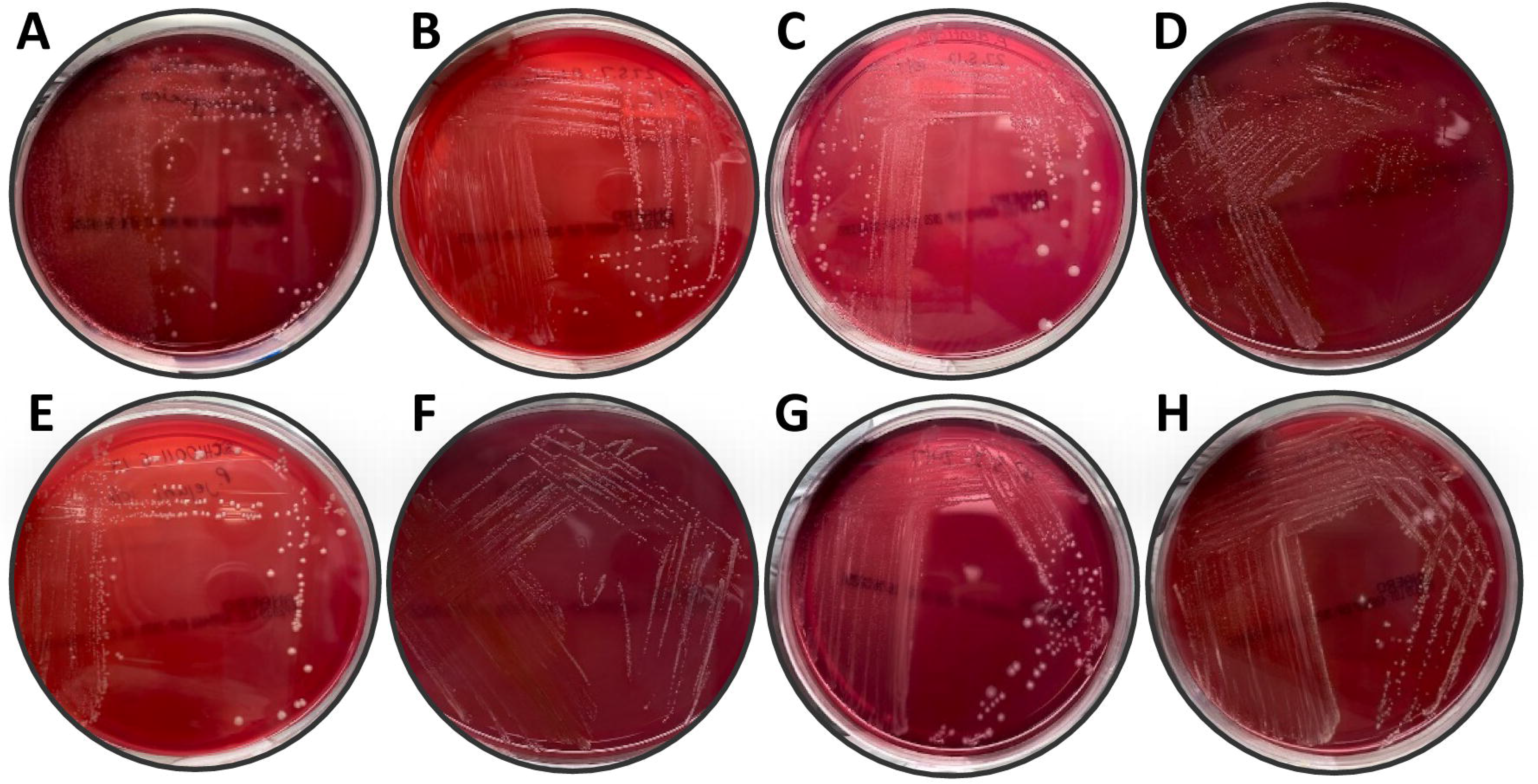
Three-day growth of *Prevotella* spp. on non-selective anaerobic blood medium. (A) *P. melaninogenica*; (B) *P. histicola*; (C) *P. denticola*; (D) *P. nanceiensis*; (E) *P. jejuni*; (F) *P. nigrescens*; (G) *P. salivae*; and (H) *P. buccae*.

**Figure 5.**
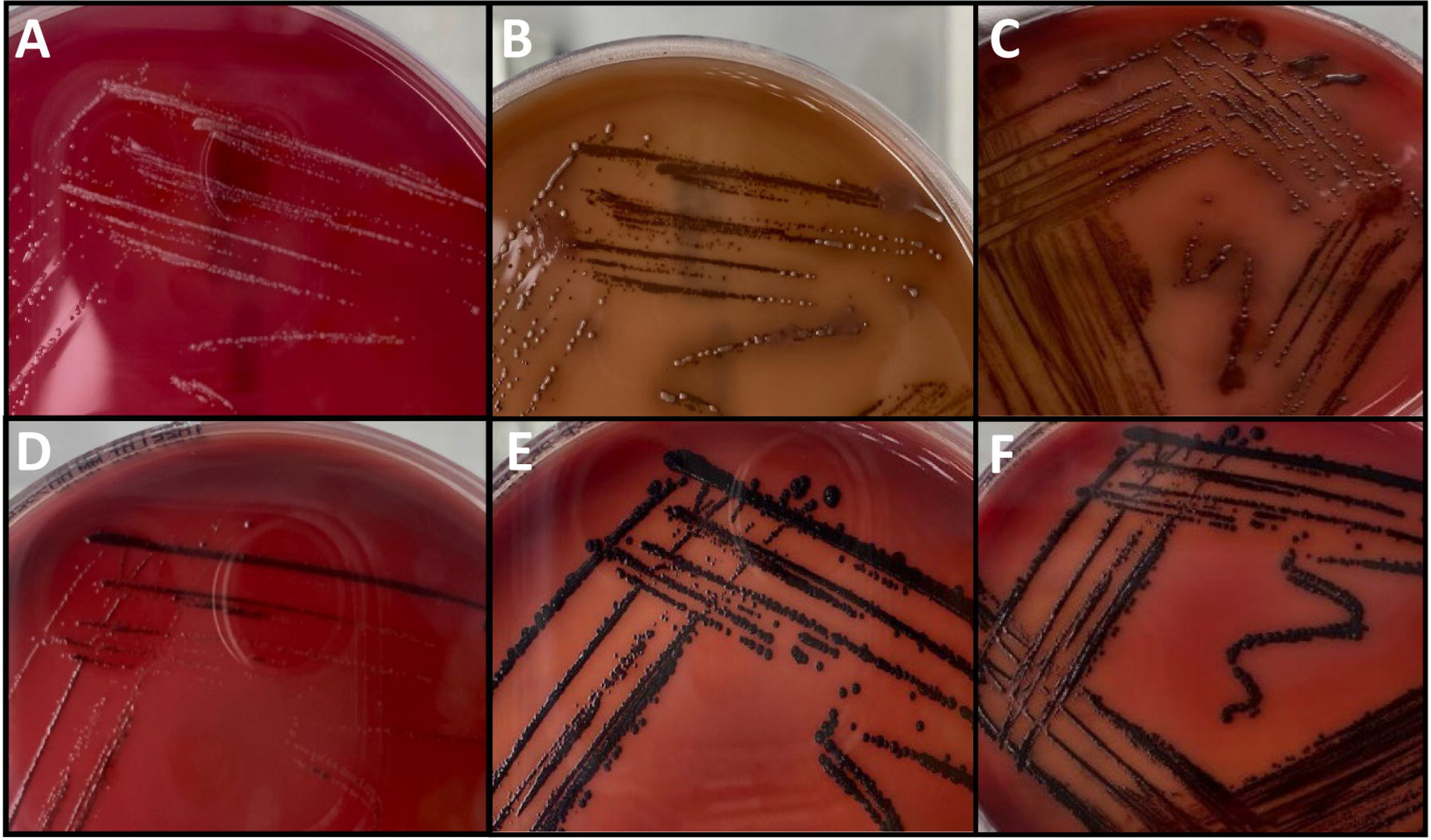
Phenotype comparison of *Prevotella nigrescens* on non-selective anaerobic blood medium (top) and polyvitaminic-supplemented chocolate agar (bottom) at (A and D) 3 days; (B and E) 5 days; and (C and F) 10 days.

**Figure 6.**
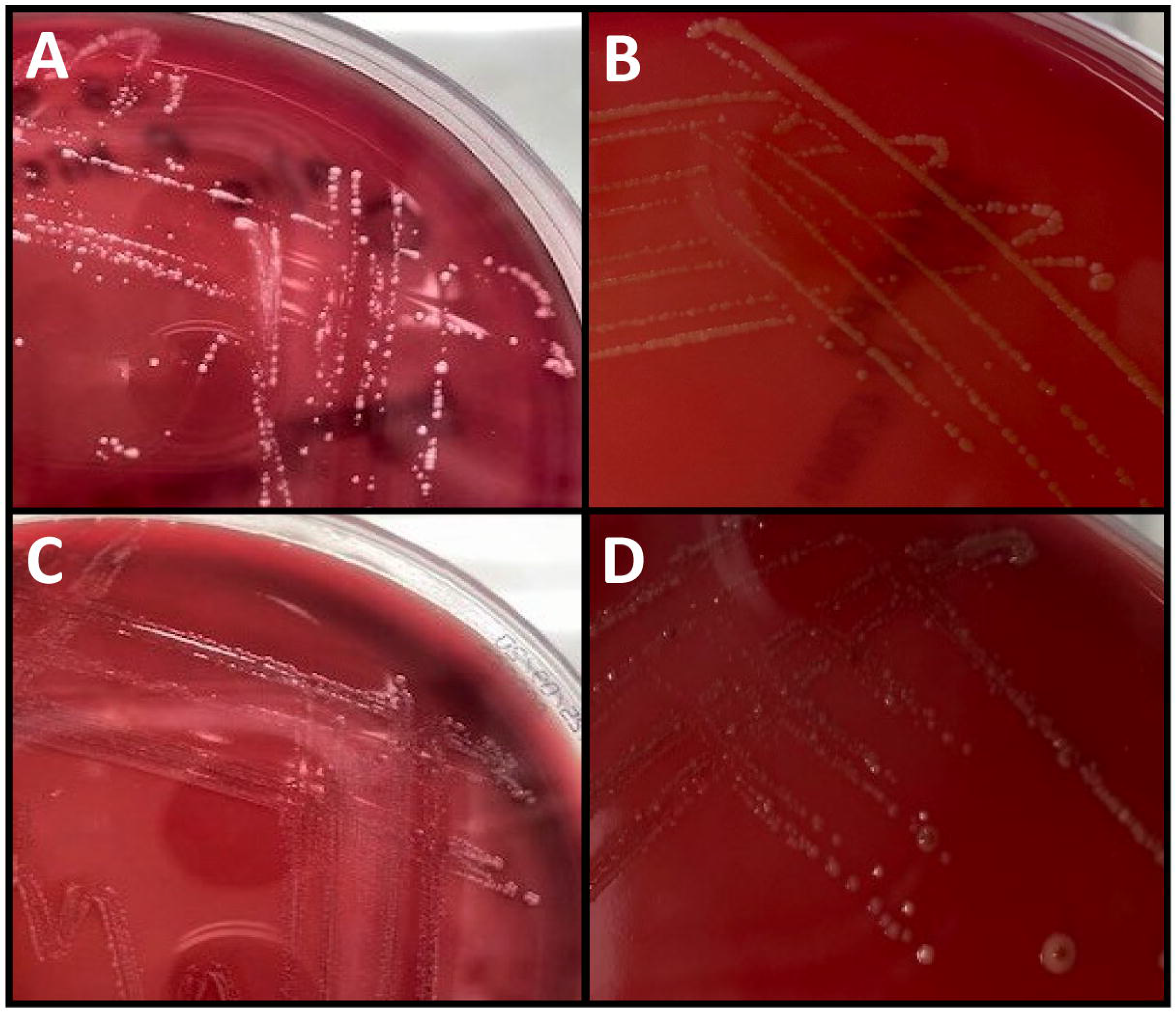
Phenotype comparison of *Prevotella histicola* on non-selective anaerobic blood medium (top) and polyvitaminic-supplemented chocolate agar (bottom) at (A and C) 3 days and (B and D) 5 days.

We acknowledge several study limitations. First, small sample size (43 enrolled participants; 69 samples) and inclusion of only COPD and CF samples makes it difficult to generalise our results across a larger cohorts. Nevertheless, the broad agreement of our findings with previous literature^10,14,19^ demonstrates a reasonable estimation of species diversity in these cohorts. Second, due to non-anaerobic sample handling and storage, non-aerotolerant *Prevotella* species may have been rendered non-viable and thus not recoverable^45^. Future establishment of disc diffusion guidelines in *Prevotella* will enable more accurate assessment of AMR phenotypes in this genus. Third, due to our use of a strict AMR cut-off (i.e. 6mm), AMR rates in our *Prevotella* are almost certainly underestimated. Fourth, we did not assess healthy airway microbiota due to difficulties associated with obtaining respiratory secretions from healthy participants. Future work should continue to seek strain-level resolution of *Prevotella* strains in CF and COPD airways, including the design of species-specific assays towards commonly isolated and known pathogenic species, assessment of *Prevotella* from healthy airways, and incorporation of higher-resolution molecular technologies such as metagenomics and metatranscriptomics to gain a clearer understanding of *Prevotella* function within complex microbiomes. Co-culture experiments between *Prevotella* species and known respiratory pathogens, such as *P. aeruginosa*, would be beneficial for elucidating the role of *Prevotella* in potentiating or abrogating pathogen virulence. Finally, phylogeographic analysis of *Prevotella* genomes may permit a deeper understanding of *Prevotella* spp. distribution and strain diversity in chronic lung disease globally, and population genomic approaches will yield a better understanding of *Prevotella* origin and capacity for airway-to-airway transmission.

## Supporting information

Merged supplemental files

## Data Availability

Thirty-five Prevotella spp. genomes generated in this study are available in the Sequence Read Archive (SRA) and GenBank databases under BioProject accession PRJNA742126.

https://www.ncbi.nlm.nih.gov/bioproject/?term=PRJNA742126

## Acknowledgements

We thank Tamieka Fraser (University of the Sunshine Coast) for laboratory assistance, Peter Timms (University of the Sunshine Coast) and Laura Sherrard (Queen’s University Belfast) for insightful feedback on this manuscript, Tania Duarte (University of Queensland) for helpful discussions about the challenges of *Prevotella* identification using current metagenomic pipelines, and the Belgian Co-ordinated Collection of Micro-organisms (BCCM) for providing type strains. This work was funded by Advance Queensland (awards AQRF13016-17RD2 and AQIRF0362018), the Wishlist Sunshine Coast Hospital Foundation (award 2019-24), and the University of the Sunshine Coast. The authors declare no conflicts of interest.

