## Supplementary material for "Genomic diversity and antimicrobial resistance of *Prevotella* spp. isolated from chronic lung disease airways": Merged supplemental files

**Table S1.** Details of cystic fibrosis (CF) and chronic obstructive pulmonary disease (COPD) samples and treatment history

| **Sample ID** | **Sample type** | **Clinical and treatment details** |
| --- | --- | --- |
| *CF-derived isolates* | | |
| SCHI0001.C.1 | Sputum | Day 10 of IV ATM and TOB |
| SCHI0002.C.1 | Sputum | Day 7 of IV CAZ, MEM, and TOB |
| SCHI0002.C.2 | Sputum | Day 37 of IV CAZ, MEM, and TOB; taken 329 days after SCHI0002.C.1 |
| SCHI0003.C.1 | Sputum | Day 4 of IV ATM, MEM, and TOB |
| SCHI0004.C.1 | Sputum | Day 14 of IV MEM and TOB |
| SCHI0005.C.1 | Sputum | Day 10 of IV MEM and TOB |
| SCHI0006.C.1 | Sputum | Clinically stable |
| SCHI0007.C.1 | Sputum | Day 3 of IV MEM and TOB |
| SCHI0008.C.1 | Sputum | Day 14 of IV ATM, CST, FOF |
| SCHI0009.C.1 | Sputum | Day 13 of FOF and TOB |
| SCHI0010.C.1 | Sputum | Day 7 of IV CAZ and TOB |
| SCHI0011.C.1 | Sputum | Day 4 of IV CAZ and TOB |
| SCHI0013.C.1 | Sputum | Day 4 of FOF and MEM |
| SCHI0014.C.1 | Sputum | Day 4 of PTZ and TOB |
| SCHI0017.C.1 | Sputum | Severe CF lung disease; Day 0; obtained just prior to antibiotic commencement |
| SCHI0018.C.1 | Sputum | Severe CF lung disease; Day 8 of IV CAZ, TZP, TOB |
| SCHI0019.C.1 | Sputum | Day 1 of IV MEM and TOB |
| SCHI0019.C.5 | Sputum | Day 11 of IV MEM and TOB |
| SCHI0019.C.10 | Sputum | Day 46; obtained 32 days after IV antibiotic cessation |
| SCHI0020.C.1 | Sputum | Day 1 of IV CAZ and TOB |
| SCHI0020.C.5 | Sputum | Day 6 of IV CAZ and TOB |
| SCHI0020.C.10 | Sputum | Day 25; obtained 16 days after IV antibiotic cessation |
| SCHI0021.C.1 | Sputum | Day 1 of IV CAZ and TOB |
| SCHI0021.C.5 | Sputum | Day 5 of IV CAZ and TOB |
| SCHI0021.C.9 | Sputum | Day 13 of IV CAZ and TOB |
| SCHI0021.C.12 | Sputum | Day 90; obtained 74 days after IV antibiotic cessation |
| SCHI0027.C.1 | Sputum | Day 1 of IV CAZ and TOB |
| SCHI0027.C.4 | Sputum | Day 9 of IV CAZ and TOB |
| SCHI0027.C.5 | Sputum | Day 12 of IV CAZ and TOB |
| SCHI0028.C.1 | Sputum | Day 0; obtained just prior to antibiotic commencement |
| SCHI0028.C.3 | Sputum | Day 7 of IV MEM and TOB |
| SCHI0028.C.5 | Sputum | Day 14 of IV MEM and TOB |
| SCHI0028.C.9 | Sputum | Day 28; obtained 13 days after IV antibiotic cessation |
| SCHI0030.C.1 | Sputum | Day 0; obtained just prior to antibiotic commencement |
| SCHI0030.C.3 | Sputum | Day 6 of IV ATM and TOB |
| SCHI0030.C.7 | Sputum | Day 13 of IV ATM and TOB |
| *COPD-derived isolates* | | |
| SCHI0034.C.1 | BW (T) | Lung cancer; collected during bronchoscopy |
| SCHI0034.C.3 | BW (Contra) | Lung cancer; collected during bronchoscopy |
| SCHI0034.C.5 | Sputum | Lung cancer; collected during bronchoscopy |
| SCHI0035.C.1 | Sputum | Stable disease |
| SCHI0036.C.1 | BW (T) | Lung cancer; collected during bronchoscopy |
| SCHI0036.C.3 | BW (Contra) | Lung cancer; collected during bronchoscopy |
| SCHI0038.C.1 | Sputum | Stable disease |
| SCHI0038.C.3 | Sputum | Stable disease |
| SCHI0039.C.1 | Sputum | Exacerbation |
| SCHI0042.C.1 | Sputum | Exacerbation; Day 7 of AMX |
| SCHI0042.C.3 | Sputum | Exacerbation but nil treatment; taken 96 days after SCHI0042.C.1 |
| SCHI0043.C.1 | BW (T) | Lung cancer; collected during bronchoscopy |
| SCHI0043.C.3 | BW (Contra) | Lung cancer; collected during bronchoscopy |
| SCHI0044.C.1 | BW (T) | Lung cancer; collected during bronchoscopy |
| SCHI0044.C.3 | BW (Contra) | Lung cancer; collected during bronchoscopy |
| SCHI0045.C.1 | Sputum | Exacerbation; hospitalised with IV TZP, oral AMC on discharge |
| SCHI0045.C.3 | Sputum | Exacerbation but nil treatment; taken 21 days after SCHI0045.C.1 |
| SCHI0046.C.1 | Sputum | Stable disease |
| SCHI0047.C.1 | Sputum | Exacerbation |
| SCHI0048.C.1 | Sputum | Stable disease |
| SCHI0049.C.1 | BW (T) | Lung cancer; collected during bronchoscopy |
| SCHI0049.C.3 | BW (Contra) | Lung cancer; collected during bronchoscopy |
| SCHI0050.C.1 | Sputum | Exacerbation; Day 3 of CIP |
| SCHI0050.C.3 | Sputum | Stable disease; taken 45 days after SCHI0050.C.1 |
| SCHI0051.C.1 | Sputum | Exacerbation; no antibiotic treatment |
| SCHI0053.C.1 | Sputum | Stable disease |
| SCHI0054.C.1 | Sputum | Stable disease |
| SCHI0054.C.3 | Sputum | Exacerbation; Day 4 of oral DOX; taken 12 days after SCHI0054.C.1 |
| SCHI0055.C.1 | BW (T) | Lung cancer; collected during bronchoscopy |
| SCHI0055.C.3 | BW (Contra) | Lung cancer; collected during bronchoscopy |
| SCHI0056.C.1 | Sputum | Exacerbation; Day 3 of oral AMX |
| SCHI0058.C.1 | Sputum | Unknown |
| SCHI0059.C.1 | Sputum | Exacerbation; Day 7 of oral AMX and DOX |

Abbreviations: AMC, amoxicillin-clavulanate; AMX, amoxicillin; ATM, aztreonam; BW, bronchial washing; CAZ, ceftazidime; CIP, ciprofloxacin; Contra, non-tumour contralateral lung site; CST, colistin; DOX, doxycycline; FOF, fosfomycin; IV, intravenous; MEM, meropenem; T, lung tumour site; TOB, tobramycin; TZP, piperacillin-tazobactam

**Table S2.** List of 60 *Prevotella* genomes used to design the *Prevotella* spp. real-time PCR assay

| **Species and strain** | **GenBank/SRA accession** |
| --- | --- |
| *P. albensis* DSM 11370 | SRR1796737 |
| *P. amnii* DNF00307 | SRR3136954 |
| *P. amnii* DSM 23384 | SRR892385 |
| *P. baroniae* DSM 16972 | SRR1796740 |
| *P. bivia* DNF00188 | SRR1518553 |
| *P. bivia* DNF00650 | SRR1518640 |
| *P. bryantii* C21a | SRR896054 |
| *P. bryantii* FB3001 | SRR4167831 |
| *P. buccalis* DNF00853 | SRR1518644 |
| *P. buccalis* DNF00985 | SRR1518651 |
| *P. corporis* DSM 18810 | SRR896458 |
| *P. corporis* MJR7716 | SRR2096011 |
| *P. dentalis* DSM 3688 | GCA_000220215.1 |
| *P. denticola* DNF00960 | SRR1518649 |
| *P. denticola* F0289 | NC_015311.1 |
| *P. denticola* KCOM1525 | NZ_CP032056.1 |
| *P. disiens* DNF00882 | SRR1518645 |
| *P. enoeca* F0113 | NZ_CP013195.1 |
| *P. fusca* JCM17724 | NZ_CP012074.1 |
| *P. histicola* T05-04(T) | SRR1518629 |
| *P. intermedia* 17-2 | NZ_AP014926.1 |
| *P. intermedia* ATCC 25611 | NZ_CP019300.1 |
| *P. intermedia* KCOM 1741 | NZ_CP024732.1 |
| *P. intermedia* KCOM 1933 | NZ_CP024729.1 |
| *P. intermedia* KCOM 1944 | NZ_CP024734.1 |
| *P. intermedia* KCOM 1949 | NZ_CP024727.1 |
| *P. intermedia* KCOM 2033 | NZ_CP024696.1 |
| *P. intermedia* KCOM 2734 | NZ_CP030094.1 |
| *P. intermedia* KCOM 2836 | NZ_CP024697.1 |
| *P. intermedia* KCOM 2837 | NZ_CP024723.1 |
| *P. intermedia* KCOM 2838 | NZ_CP024725.1 |
| *P. intermedia* OMA14 | NZ_AP014597.1 |
| *P. intermedia* 17 | NC_017860.1 |
| *P. jejuni* CDC3:33 | NZ_CP023863.1 |
| *P. loescheii* DSM 19665 | SRR892376 |
| *P. maculosa* DSM 19339 | SRR891726 |
| *P. melaninogenica* ADL-403 | SRR755359 |
| *P. melaninogenica* ATCC 25845 | NC_014370.1 |
| *P. melaninogenica* DNF00666 | SRR1518642 |
| *P. melaninogenica* FDAARGOS_306 | NZ_CP022040.2 |
| *P. melaninogenica* GAI07411 | NZ_AP018049.1 |
| *P. nigrescens* ATCC 33563 | AFPX01000001.1 |
| *P. oris* DSM 18711 | SRR892475 |
| *P. oris* NCTC 13071 | NZ_LR134384.1 |
| *P. ruminicola* 23 | NC_014033.1 |
| *P. saccharolytica* JCM 17484 | BAKN01000001.1 |
| *P. shahii* DSM 15611 | SRR4096625 |
| *Prevotella* sp. AGR2160 | SRR896023 |
| *Prevotella* sp. MA2016 | SRR1588129 |
| *Prevotella* sp. FD3004 | SRR1588128 |
| *Prevotella* sp. HJM029 | SRR1045121 |
| *Prevotella* sp. MSX73 | SRR445467 |
| *Prevotella* sp. NE3005 | SRR4167832 |
| *Prevotella* sp. oral taxon F0039 | GCA_000163055.2 |
| *Prevotella* sp. oral taxon F0472 | SRR387761 |
| *Prevotella* sp. S7-1-8 | SRR1518484 |
| *P. stercorea* AF42-9 | QRNO01000001.1 |
| *P. timonensis* DSM 22865 | SRR896444 |
| *P. timonensis* S9-PR14 | SRR1518578 |
| *P. veroralis* DSM 19559 | SRR892391 |

**Table S3.** Summary of *Prevotella* results from microbiological culture, real-time PCR, 16S rRNA relative abundance, and whole-genome sequencing of cystic fibrosis (CF) and chronic obstructive pulmonary disease (COPD) samples.

| Specimen ID | Specimen type | PCR result^†^ | *Prevotella* 16S %* | Culture result | Isolate/s | WGS result | WGS accession |
| --- | --- | --- | --- | --- | --- | --- | --- |
| *CF* | | | | | | | |
| SCHI0001.C.1 | Sputum | + | 24.9 | + | SCHI0001.S.7 | Mix: *P. salivae* (15%) | NA |
| SCHI0002.C.1 | Sputum | + | 6.6 | + | SCHI0002.S.13 | Not sequenced | NA |
| SCHI0002.C.2 | Sputum | + | 0.3 | + | SCHI0002.S.15 | Not sequenced | NA |
| SCHI0003.C.1 | Sputum | - | 0 | - | NA | NA | NA |
| SCHI0004.C.1 | Sputum | - | 0 | - | NA | NA | NA |
| SCHI0005.C.1 | Sputum | + | 1.3 | - | NA | NA | NA |
| SCHI0006.C.1 | Sputum | + | 6.8 | + | SCHI0006.S.13  SCHI0006.S.15  SCHI0006.S.17 | *P. histicola*  Mix: *P. buccae* (12%), *P. oralis* (5%)  Mix: *P. buccae* (46%) | JAHXCX000000000  NA  NA |
| SCHI0007.C.1 | Sputum | + | 2.1 | - | NA | NA | NA |
| SCHI0008.C.1 | Sputum | + | 48.8 | - | NA | NA | NA |
| SCHI0009.C.1 | Sputum | + | nt | + | SCHI0009.S.5  SCHI0009.S.6 | *P. melaninogenica*  *P. histicola* | JAHXCW000000000  JAHXCV000000000 |
| SCHI0010.C.1 | Sputum | + | 21.4 | + | SCHI0010.S.3 | *P. nanceiensis* | JAHXCU000000000 |
| SCHI0011.C.1 | Sputum | + | 32.6 | + | SCHI0011.S.12  SCHI0011.S.13 | *P. nanceiensis*  *P. jejuni* | JAHXCT000000000  JAHXCS000000000 |
| SCHI0013.C.1 | Sputum | - | nt | - | NA | NA | NA |
| SCHI0014.C.1 | Sputum | + | 74.3 | - | NA | NA | NA |
| SCHI0017.C.1 | Sputum | - | nt | - | NA | NA | NA |
| SCHI0018.C.1 | Sputum | + | nt | + | SCHI0018.S.9 | *P. salivae* | JAHXCR000000000 |
| SCHI0019.C.1 | Sputum | + | 1.6 | - | NA | NA | NA |
| SCHI0019.C.5 | Sputum | - | 0 | - | NA | NA | NA |
| SCHI0019.C.10 | Sputum | + | 3.3 | - | NA | NA | NA |
| SCHI0020.C.1 | Sputum | + | 12 | - | NA | NA | NA |
| SCHI0020.C.5 | Sputum | + | 41.6 | - | NA | NA | NA |
| SCHI0020.C.10 | Sputum | + | 2.8 | - | NA | NA | NA |
| SCHI0021.C.1 | Sputum | + | 10.3 | + | SCHI0021.S.9 | *P. denticola* | JAHXCQ000000000 |
| SCHI0021.C.5 | Sputum | + | 8.9 | + | SCHI0021.S.13 | Not sequenced | NA |
| SCHI0021.C.9 | Sputum | + | 14.6 | - | NA | NA | NA |
| SCHI0021.C.12 | Sputum | + | 5.6 | - | NA | NA | NA |
| SCHI0027.C.1 | Sputum | + | 29.9 | + | SCHI0027.S.6  SCHI0027.S.7  SCHI0027.S.8  SCHI0027.S.10 | *P. melaninogenica*  *P. histicola*  Mix: *P. histicola* (52%), *P. melaninogenica* (47%)  Mix: *P. salivae* (27%) | JAHXCP000000000  JAHXCO000000000  NA  NA |
| SCHI0027.C.4 | Sputum | + | 13.4 | + | SCHI0027.S.9  SCHI0027.S.11 | *P. melaninogenica*  *P. melaninogenica* | JAHXCN000000000  JAHXCM000000000 |
| SCHI0027.C.5 | Sputum | + | 3.6 | + | SCHI0027.S.12  SCHI0027.S.13  SCHI0027.S.14 | *P. denticola*  *P. melaninogenica*  *P. denticola* | JAHXCL000000000  JAHXCK000000000  JAHXCJ000000000 |
| SCHI0028.C.1 | Sputum | + | 32.8 | - | NA | NA | NA |
| SCHI0028.C.3 | Sputum | + | 6.2 | - | NA | NA | NA |
| SCHI0028.C.5 | Sputum | + | 10.9 | + | SCHI0028.S.1 | *P. histicola* | JAHXCI000000000 |
| SCHI0028.C.9 | Sputum | + | nt | + | SCHI0028.S.4  SCHI0028.S.5 | *P. histicola*  *P. nigrescens* | JAHXCH000000000  JAHXCG000000000 |
| SCHI0030.C.1 | Sputum | + | 4.2 | + | SCHI0030.S.6 | Mix: *P. salivae* (17%)*, P. melaninogenica* (1%) | NA |
| SCHI0030.C.3 | Sputum | - | 0.0 | - | NA | NA | NA |
| SCHI0030.C.7 | Sputum | - | 0.0 | - | NA | NA | NA |
| *COPD* | | | | | | | |
| SCHI0034.C.1 | BW (T) | + | nt | - | NA | NA | NA |
| SCHI0034.C.3 | BW (CL) | + | nt | + | SCHI0034.S.5  SCHI0034.S.6  SCHI0034.S.7  SCHI0034.S.8 | *P. histicola*  *P. histicola*  *P. melaninogenica*  *P. salivae* | JAHXCF000000000  JAHXRH000000000  JAHXRG000000000  JAHXQY000000000 |
| SCHI0034.C.5 | Sputum | + | nt | + | SCHI0034.S.9  SCHI0034.S.10  SCHI0034.S.11 | *P. melaninogenica*  *P. histicola*  *P. salivae* | JAHXCB000000000  JAHXRE000000000  JAHXRD000000000 |
| SCHI0035.C.1 | Sputum | - | nt | - | NA | NA | NA |
| SCHI0036.C.1 | BW (T) | - | 0.004 | - | NA | NA | NA |
| SCHI0036.C.3 | BW (CL) | - | 0.009 | - | NA | NA | NA |
| SCHI0038.C.1 | Sputum | - | nt | - | NA | NA | NA |
| SCHI0038.C.3 | Sputum | - | nt | - | NA | NA | NA |
| SCHI0039.C.1 | Sputum | - | 0.3 | - | NA | NA | NA |
| SCHI0042.C.1 | Sputum | + | 1.2 | + | SCHI0042.S.4  SCHI0042.S.5 | *P. denticola*  *P. nanceiensis* | JAHXRC000000000  JAHXRB000000000 |
| SCHI0042.C.3 | Sputum | + | nt | + | SCHI0042.S.6  SCHI0042.S.7 | Not sequenced  Not sequenced | NA  NA |
| SCHI0043.C.1 | BW (T) | + | 25.7 | + | SCHI0043.S.1  SCHI0043.S.2 | *P. melaninogenica*  *P. melaninogenica* | JAHXRA000000000  JAHXCD000000000 |
| SCHI0043.C.3 | BW (CL) | + | nt | + | SCHI0043.S.3 | *P. melaninogenica* | JAHXCC000000000 |
| SCHI0044.C.3 | BW (CL) | + | 0.4 | + | SCHI0044.S.1 | *P. melaninogenica* | JAHXQZ000000000 |
| SCHI0045.C.1 | Sputum | - | 0.09 | - | NA | NA | NA |
| SCHI0045.C.3 | Sputum | + | 0.4 | - | NA | NA | NA |
| SCHI0046.C.1 | Sputum | + | 3.2 | - | NA | NA | NA |
| SCHI0047.C.1 | Sputum | + | 36.4 | + | SCHI0047.S.2  SCHI0047.S.3  SCHI0047.S.4 | *P. melaninogenica*  *P. salivae*  *P. buccae* | JAHXCE000000000  JAHXRF000000000  JAHXQX000000000 |
| SCHI0048.C.1 | Sputum | - | 0.02 | - | NA | NA | NA |
| SCHI0049.C.1 | BW (T) | - | 0.07 | - | NA | NA | NA |
| SCHI0049.C.3 | BW (CL) | + | 0.00 | + | SCHI0049.S.1 | *P. melaninogenica* | JAHXQW000000000 |
| SCHI0050.C.1 | Sputum | - | 0.00 | - | NA | NA | NA |
| SCHI0050.C.3 | Sputum | - | NA | - | NA | NA | NA |
| SCHI0051.C.1 | Sputum | + | 12.7 | - | NA | NA | NA |
| SCHI0053.C.1 | Sputum | + | 0.08 | - | NA | NA | NA |
| SCHI0054.C.1 | Sputum | + | 16.0 | - | NA | NA | NA |
| SCHI0054.C.3 | Sputum | + | 3.0 | - | NA | NA | NA |
| SCHI0055.C.1 | BW (T) | + | 0.17 | - | NA | NA | NA |
| SCHI0055.C.3 | BW (CL) | + | 0.8 | - | NA | NA | NA |
| SCHI0056.C.1 | Sputum | + | 2.8 | - | NA | NA | NA |
| SCHI0058.C.1 | Sputum | + | NA | - | NA | NA | NA |
| SCHI0059.C.1 | Sputum | + | 1.1 | - | NA | NA | NA |

^†^On total DNA extracted from the clinical specimen. BW (T), Bronchial washing from tumour site; BW (CL), Bronchial washing from contralateral lung

*Relative *Prevotella* abundance according to 16S rRNA gene metataxonomics

#According to Ribosomal MLST (Jolley *et al*., 2012 *Microbiology (Reading)*:158(Pt 4):1005-1015.

nt, not tested; NA, not applicable

**^**No DNA was available for testing of the total specimen
